# Chest CT Images for COVID-19: Radiologists and Computer-based detection

**DOI:** 10.1101/2020.06.27.20141531

**Authors:** Qingli Dou, Jiangping Liu, Wenwu Zhang, Yanan Gu, Wan-Ting Hsu, Kuan-Ching Ho, Hoi Sin Tong, Wing Yan Yu, Chien-Chang Lee

## Abstract

**Background:** Characteristic chest computed tomography (CT) manifestation of 2019 novel coronavirus (COVID-19) was added as a diagnostic criterion in the Chinese National COVID-19 management guideline. Whether the characteristic findings of Chest CT could differentiate confirmed COVID-19 cases from other positive nucleic acid test (NAT)-negative patients has not been rigorously evaluated.

**Purpose:** We aim to test whether chest computed tomography (CT) manifestation of 2019 novel coronavirus (COVID-19) can be differentiated by a radiologist or a computer-based CT image analysis system.

**Methods:** We conducted a retrospective case-control study that included 52 laboratory-confirmed COVID-19 patients and 80 non-COVID-19 viral pneumonia patients between 20 December, 2019 and 10 February, 2020. The chest CT images were evaluated by radiologists in a double blind fashion. A computer-based image analysis system (uAI system, Lianying Inc., Shanghai, China) detected the lesions in 18 lung segments defined by Boyden classification system and calculated the infected volume in each segment. The number and volume of lesions detected by radiologist and computer system was compared with Chi-square test or Mann-Whitney U test as appropriate.

**Results:** The main CT manifestations of COVID-19 were multi-lobar/segmental peripheral ground-glass opacities and patchy air space infiltrates. The case and control groups were similar in demographics, comorbidity, and clinical manifestations. There was no significant difference in eight radiologist identified CT image features between the two groups of patients. There was also no difference in the absolute and relative volume of infected regions in each lung segment.

**Conclusions:** We documented the non-differentiating nature of initial chest CT image between COVID-19 and other viral pneumonia with suspected symptoms. Our results do not support CT findings replacing microbiological diagnosis as a critical criterion for COVID-19 diagnosis. Our findings may prompt re-evaluation of isolated patients without laboratory confirmation.

## INTRODUCTION

Due to high transmissibility and so far lack of proven treatment, the 2019 novel coronavirus disease, 2019-nCoV, has quickly disseminated worldwide.^4,5^ As symptoms of COVID-19 are similar to other acute respiratory infections, diagnosis relies on positive nucleic acid test (NAT). Given the long turnaround time and suboptimal sensitivity of NAT, chest computed tomography (CT) was proposed as a first line diagnostic tool by the Chinese national guideline (trial version 5).^7^ After the new definition implementation, 14,840 new cases with 242 deaths were reported on February 13th, which was the record by far reported in a single day since the outbreak.

Several case series have reported characteristic CT findings of COVID-19, including ground glass opacities in bilateral peripheral lung, crazy-paving changes, reticular thickening, or consolidations.^8,9^We aim to evaluate whether radiologists or a computer-based image analysis system can reliably differentiate COVID-19 cases from non-COVID-19 but suspected patients.

## METHODS

### Study population

From 20 December, 2019 to 10 February, 2020, 52 laboratory-confirmed COVID-19 patients in Shenzhen were identified that fulfilled the diagnostic criteria of Chinese national guideline.^10^ The criteria for a confirmed case include documented laboratory evidence, compatible clinical symptoms, and exposure history. Documented laboratory evidence is defined by positive NAT result either from respiratory tract, bronchoalveolar lavage fluid, or blood sample. Compatible clinical symptoms refer to fever, cough, imaging characteristics of pneumonia, and/or normal or decreased white blood cells count or decreased lymphocyte count. Exposure history includes travel/residence history in Wuhan city, contact history with laboratory-confirmed patients, or contact history with patients with fever or respiratory symptoms from Wuhan and its surrounding areas or endemic communities, within 14 days before the onset of illness. We randomly selected 80 laboratory-confirmed non-COVID-19 viral pneumonia patients as controls. These presented with suspected symptoms and exposure history and underwent NAT and chest CT exams in the same period. These patients must have at least two negative NAT results and a laboratory evidence of other respiratory virus infection.

### Chest CT evaluation by radiologists

All CT images were independently retrospectively analyzed using a structured form by two experienced radiologists in a double blinded fashion without knowing the clinical diagnosis. A third senior radiologist was consulted to solve any discrepancy by consensus. Evaluation was focused on the presence of ground glass opacities, patchy infiltration, patchy consolidation, pleural effusion, mediastinal lymphadenopathy, air bronchogram, pleural thickening, and Interstitial change.

### Machine-learning based CT lesion detection and quantification system

We used the uAI image analysis system (Lianying Intelligent Medical Technology Co Ltd, Shanghai, China) for detection and quantification of chest CT lesions (Computer-based detection in Appendix). The system classifies the lung fields into five lung lobes and 18 lung segments based on Boyden classification, detected infected regions in each anatomical region, and quantified the cumulative and relative infected volume. The infected volumes between the two groups were also compared in four different CT windows. This study was approved by the Institutional Review Board of the Second Affiliated Hospital of Shenzhen University.

### Statistical analysis

Categorical variables were expressed as number and proportion and compared with a Chi-square test. Continuous variables were presented with mean ± standard deviation for data with a normal distribution and tested by Student t test. Data with non-normal distribution were presented with median with interquartile range and compared with independent sample Mann-Whitney U test. All tests in this study were two-sided, and P <0.05 was deemed statistically significant. Data were analyzed using SPSS 23.0 software.

## RESULTS

### Study patients

There was no significant difference between case and control cases in all categories (Table 1).

**Table 1.** Patients characteristics between case and control patients [n(%)]

| Characteristic | COVID-19<br>(N=52) | non-COVID-19 viral<br>pneumonia<br>(N =80) | P Value |
| --- | --- | --- | --- |
| Age | 45.61±14.19 | 48.69±14.62 | 0.12 |
| Sex |  |  |  |
| Female | 32(61.54%) | 58(72.50%) | 0.44 |
| Male | 20(38.46%) | 22(27.50%) | 0.73 |
| Course of disease | 5.61±2.19 | 8.32±1.82 | 0.61 |
| Onset of symptom to CT<br>examination | 4.32±0.82 | 6.87±1.89 | 0.06 |
| Symptoms |  |  |  |
| Fever | 47(90.38) | 71(88.75) | 0.24 |
| Fatigue | 44(84.61) | 68(85.00) | 0.08 |
| Dry cough | 31(59.62) | 48(60.00) | 0.16 |
| Dyspnea | 15(28.85) | 27(33.75) | 0.67 |
| Sore throat | 28(53.85) | 49(61.25) | 0.48 |
| Comorbidities |  |  |  |
| Hypertension | 24(46.15) | 39(48.75) | 0.89 |
| Chronic heart failure | 10(19.23) | 17(21.25) | 0.85 |
| Diabetes | 14(26.92) | 20(25.00) | 0.52 |
| Cerebrovascular disease | 9(17.31) | 19(23.75) | 0.37 |
| COPD | 11(21.15) | 16(20.00) | 0.94 |
| Chronic kidney disease | 4(7.69) | 8(10.00) | 0.30 |
| Laboratory results |  |  |  |
| White blood cell count (10<sup>9</sup>/L) | 13.60±5.38 | 11.35±5.88 | 0.39 |
| Neutrophil percentage (%) | 20.43±5.86 | 18.20±5.05 | 0.59 |
| Lymphocyte percentage (%) | 82.45±14.54 | 85.69±10.08 | 0.20 |
| C-reactive protein (mg/dL) | 93.20±24.01 | 106.59±29.29 | 0.10 |
| Lactate (mmol/L) | 2.71±1.38 | 2.58±1.20 | 0.16 |

### Comparison of CT interpretation by radiologists

In both COVID-19 and non-COVID-19 viral pneumonia patients, ground-glass opacity (Supplemental Figures 1A and 1B) and patchy airspace infiltrates (Supplemental Figures 2A and 2B) were the major findings. The lesions could be found in multiple lobes or segments, more often bilateral. There was no statistical significance between the distribution of lesions identified by radiologists in the two groups (Table 2).

**Table 2.** Radiologist interpretation of chest CT before NAT results [n(%)]

|  | COVID-19<br>(N=52) | non-COVID-19 viral<br>pneumonia (N =80) | *P* Value |
| --- | --- | --- | --- |
| Ground glass opacities | 38(73.07) | 57(71.25) | 0.27 |
| Patch infiltration | 30(57.69) | 48(60.00) | 0.08 |
| Patch consolidation | 15(28.84) | 28(35.00) | 0.15 |
| Pleural effusion | 4(7.69) | 9(11.25) | 0.54 |
| Mediastinal lymphadenopathy | 3(5.78) | 5(6.25) | 0.71 |
| Air bronchogram | 11(21.15) | 15(18.75) | 0.19 |
| Pleural thickening | 4(7.69) | 6(7.50) | 0.57 |
| Interstitial change | 10(19.23) | 16(20.00) | 0.29 |

### Comparison of computer system detected infected lung volume

Compared between COVID-19 and non-COVID-19 viral pneumonia patients, there was no significant difference in the computer system detected infected lung volume /percentage in 5 lung lobes and in different lung segments (Table 3). Similarly, there was no significant difference in the absolute or relative infected lung volume between the two groups of patients (Table S1).

**Table 3.**
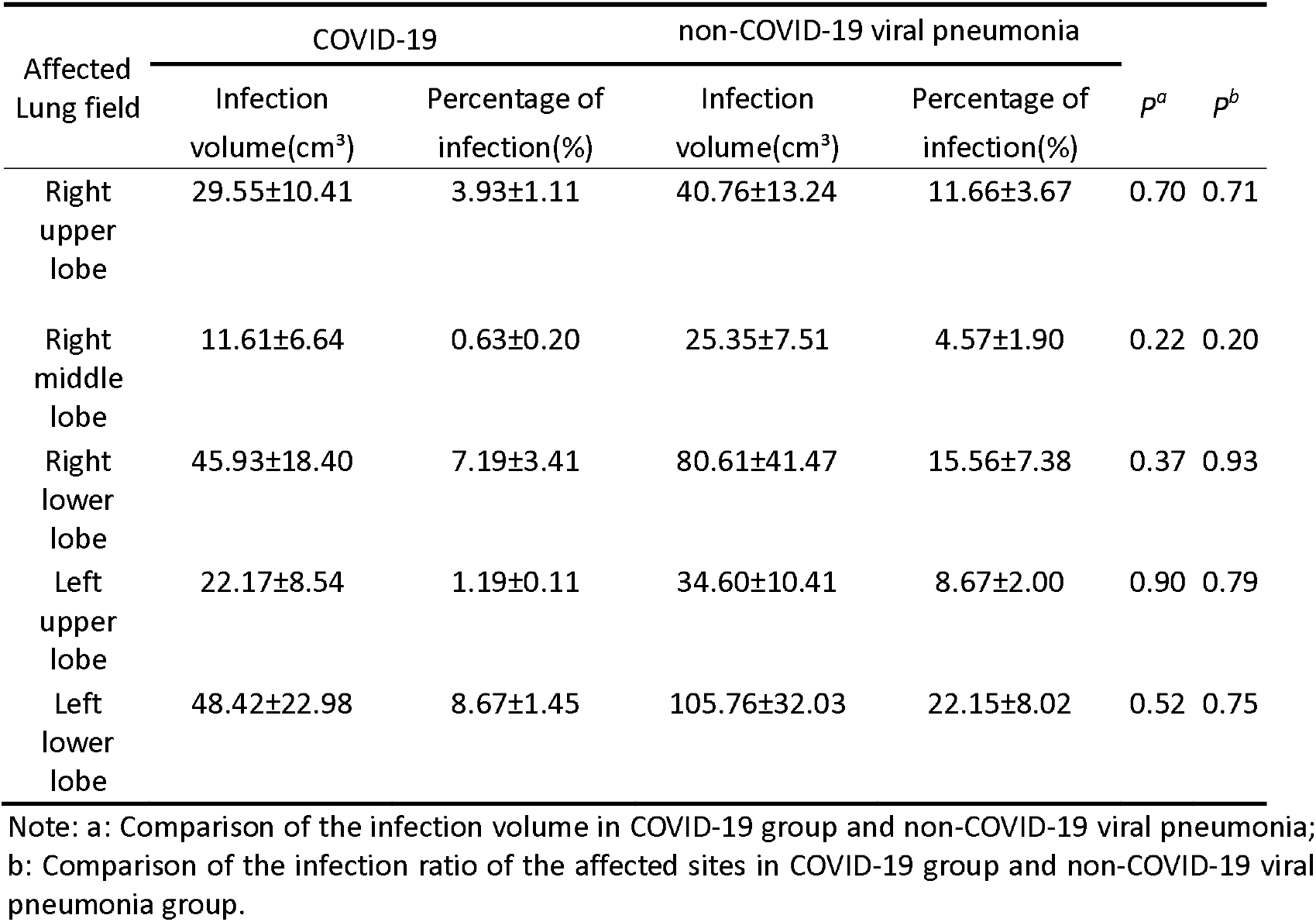
Difference in lesions distribution between COVID-19 and non-COVID-19 viral pneumonia patients.

### Sensitivity analysis under different radiodensity windows for machine detection

The lesions’ infection volume/percentage for COVID-19 and non-COVID-19 patients were stratified by four different CT number ranges (Table S2). Lesions in the [-750, -300) Hounsfield units (HU) range were arbitrarily chosen to reflect ground-glass opacity, while those with CT numbers in the [-300, 50) range were considered as denser airspace infiltrates and consolidation. Anything less than -750 becomes harder to differentiate from normal lung tissue. An extreme number 50 was chosen, beyond which infection becomes unlikely, though not impossible, as the number starts to get into the soft tissue mass range. No statistically significant difference in the infected volumes between the two groups was found in any of the four ranges.

## DISCUSSION

Chen et al. studied 29 patients with COVID-19 showed that the chest image lesions were mostly bilateral and multiple with patchy shadows and ground glass opacities.^11^.^12^ Pan et al reported CT images of COVID-19 are diverse in the early stages, which may present ground-glass opacities, pulmonary consolidation and nodules.^13^These studies were case series without suitable control groups.^8,9,13-15^ We found Radiologists’ interpretation alone or computer-based lesion detection cannot differentiate COVID-19 form other viral pneumonia. Such findings were robust under different anatomic sites or CT density ranges.

There are pros and cons using clinical diagnosis as a case definition. Clinical diagnosis allows early isolation with initial false negative NAT, slows transmission, and implements treatment early. However, the use of chest CT as a first line diagnostic method may miss early/mild disease, promote cross-infection in the CT room, increase radiation exposure, and consume enormous resources of disinfection.

Our findings have multiple implications. First, clinicians should not rely on initial chest CT findings to diagnose COVID-19 in the absence of laboratory confirmation. Second, patients who had been diagnosed with COVID-19 based on the clinical grounds should be re-evaluated with serum antibody tests. Third, patients who were isolated and cohorted with laboratory-confirmed cases in the temporary COVID-19 hospitals of Hubei province may need to be reevaluated aiming for further laboratory evidence including repeated NAT or serum antibody test.

Results of study should be interpreted in light of its shortfalls. Due to the constraints in time and patient number, we could not perform independent validation with an external sample. We did not compare serial imaging changes. We used a commercial image analysis system based on deep learning. The system does not provide flexibility in adjustment of machine-learning model or hyperparameters selection. We do not exclude future tailor trained machine learning systems that can differentiate chest CT of COVID-19 from other suspected patients.

## CONCLUSION

We documented the non-differentiating nature of initial chest CT between COVID-19 and other viral pneumonia with suspected symptoms. Our results do not support CT findings replacing microbiological diagnosis as a critical criterion for COVID-19 diagnosis. Our findings may prompt re-evaluation of isolated patients without laboratory confirmation.

## Data Availability

The data sets generated and/or analyzed during the current study are not publicly available due to the data confidentiality requirements of the ethics committee, but are available from the corresponding author on reasonable request and approval from the ethics committees in all institutions.

## Author Contributions

Conceptualization: Qingli Dou, Jiangping Liu. Data curation: Qingli Dou, Jiangping Liu. Formal analysis: Qingli Dou, Jiangping Liu.

Methodology: Qingli Dou, Jiangping Liu, Wenwu Zhang, Yanan Gu Project administration: Wan-Ting Hsu, Hoi Sin Tong, Wing Yan Yu. Resources: Qingli Dou, Jiangping Liu, Chien-Chang Lee. Supervision: Qingli Dou, Chien-Chang Lee.

Writing – original draft: Qingli Dou, Jiangping Liu, Wenwu Zhang, Yanan Gu, Chien-Chang Lee.

Writing – review & editing: Qingli Dou, Jiangping Liu, Kuan-Ching Ho, Hoi Sin Tong, Wing Yan Yu, Chien-Chang Lee.

## Acknowledgement

We are indebted to Dr. Anuj Pareek at Department of Radiology, Stanford University for his constructive comments on the manuscript.

